# Teaching medical students about Kawasaki Disease: The development and evaluation of a digital educational resource

**DOI:** 10.1101/2022.10.01.22280590

**Authors:** Jong Eun Song, Michal Tombs

**Author notes:** Corresponding author: Jong Eun Song.

## Abstract

**Introduction:** The use of digital educational resources has gained widespread popularity across all disciplines in medical education; paediatrics being no exception. This paper focuses on the development and in particular, the evaluation, of an e-learning resource on Kawasaki Disease which was primarily developed as a revision aid for undergraduate medical students.

**Methods:** This digital resource was created using Xerte Online toolkit and was guided by the instructional model ADDIE. The design and development of the resource was based on Mayer’s 12 Multimedia Principles, while Zaharias and Poylymenakou’s usability evaluation method informed the preliminary evaluation.

**Findings:** Feedback from the preliminary evaluation, which was based on the instructional design parameters of ‘navigation’, ‘visual design’ and ‘intrinsic motivation to learn’, was overwhelmingly positive. However, in light of this being a small-scale evaluation conducted on just seven undergraduate students, a protocol was developed for a more in-depth and robust evaluation.

**Conclusions and further development:** The evaluation of digital educational resources remains a fundamental and essential component in creating meaningful and effective e-learning material. In line with literature on evaluation of digital educational resources in medical education, the evaluation protocol set out in this paper considers three aspects; evaluating the ability of the resource to meet its intended outcomes; evaluating the ability of the resource to meet learners’ needs; and evaluating the educational context.

## Introduction

Medical educators increasingly utilise digital educational resources to disseminate content and knowledge in their teaching (Huynh 2017). These resources are used either as a standalone learning tool or as part of ‘flipped learning’; a pedagogical approach that allows students to acquire foundational knowledge during independent study, which can then be applied and expanded on in the classroom (Moffett 2015). In line with adult learning theory, this shifts the focus from educator to learner-centred learning (Ruiz et al. 2006), with research showing the benefits of the approach for improving knowledge retention and learner engagement. Indeed, evidence suggests that engaging with such resources is a more efficient way of acquiring knowledge and skills compared to traditional educator-led teaching, which in turn can lead to increased motivation among learners (Cullen et al. 2019).

Within paediatric education, digital educational resources covering a wide range of essential skills have been created globally. While some resources are designed for the purposes of providing quick access to high-yield information such as the Peds Cardiology Handbook app (Rochelson et al. 2022), others are targeted towards a wider audience and take the form of written, audio and video content, as discussed in Martiniuk et al’s study (2022). Resources have also been created to provide training in areas of paediatrics that traditionally take place face-to-face, such as paediatric resuscitation (O’Leary 2012) and communication (Kranenburg et al. 2017). Both studies found this mode of teaching to be effective in improving knowledge and competence in their respective areas among healthcare professionals.

It is worth mentioning that digital educational resources are not always developed by expert content creators; for example, Peds Cases is a highly successful student-driven, peer reviewed interactive website comprising of quizzes, cases and clinical videos that is aligned to the paediatric undergraduate medical education curriculum at the University of Alberta (Gill et al. 2010). Therefore, it is possible for medical educators with initiative and sound educational principles to create content that is enriching and beneficial for learners across the globe.

Tools to create digital resources are readily available and accessible for teaching clinicians; an example being Xerte, which is a free, open-source software (University of Nottingham, 2022). Several Xerte e-learning resources, primarily focusing on design and development, have been published in literature by medical educators (Salmon et al. 2019; Sait and Tombs 2021). However given the proliferation of digital educational resources in the last couple of decades (Leacock and Nesbit 2007), there remains a need to focus our attention on the evaluation of such tools to enhance e-learning quality and user satisfaction. This paper will hone in on the preliminary evaluation of a Xerte e-learning resource on Kawasaki Disease, which was created primarily as a revision aid for medical students. Following this, an evaluation protocol on performing a more robust evaluation of the Xerte resource will be presented. The Xerte resource on Kawasaki Disease can be accessed using the following link: https://xerte.cardiff.ac.uk/play_14952.

## Methods

### The development of the resource

The Analysis, Design, Development, Implementation and Evaluation (ADDIE) instructional model guided the development of this resource (Khalil and Elkhider 2016). In the initial stage, a PACT (People, Activities, Contexts and Technologies) analysis was conducted on the target audience to explore learners’ needs and requirements of the resource. This took place through informal discussions with medical students via common social media platforms. Analysis revealed that students were in favour of utilising digital resources for revision purposes, as accessing educational content via hand-held devices such as mobile phones and tablets offers great flexibility. The interactivity of a digital resource was highlighted as an appealing feature; the overriding opinion being that this would serve as an engaging and enjoyable alternative to revising from textbooks. This is in keeping with literature which suggests that many of the ‘generation Y’ students have a preference for e-learning packages that have an interactive component to it over textbooks (McKendree 2010). Students were unanimous in that the topic for the digital resource would have to be both important and relevant to their learning.

Kawasaki Disease is an acute, self-limited vasculitis of childhood that is the most common cause of cardiovascular disease in childhood. Prompt diagnosis is crucial as a delay in treatment with high dose intravenous immunoglobulins can lead to one in five children developing coronary artery aneurysms (Newburger Jane et al. 2016). Owing to nonspecific clinical signs and the lack of a definitive diagnostic test, it can be a difficult condition to diagnose. However, there is a criteria established by Tomisaku Kawasaki (1967) that clinicians refer to, and which medical students are expected to learn about through independent study and paediatric clinical attachments. Given that the incidence of Kawasaki Disease stands at around 8 in 100,000 in the UK (NHS 2021), the chances of medical students encountering a patient with this condition are scarce. Further, during the COVID-19 pandemic, clinical placements came to a halt, which left students with no option but to turn to books and alternative resources to supplement their learning. This digital resource was therefore developed as an interactive revision aid that students could undertake in their own time and at their own pace. Following the PACT analysis, specific learning outcomes were developed in the Design phase using measurable verbs such as ‘explain’ and ‘describe’ (see Box 1). These learning outcomes were based upon the lower-order skills in Bloom’s taxonomy (Forehand 2010).

#### Box 1

Learning outcomes of the digital educational resource

By the end of this session, you should be able to:

- Explain the epidemiology, aetiology, diagnostic criteria, and the treatment options for Kawasaki Disease
- List the differential diagnosis for Kawasaki Disease
- Describe the common complications of Kawasaki Disease

In the Development stage, Mayer’s multimedia principles were applied (Mayer 2010). Based on cognitive load theory, the underpinning assumption is that an individual has limited cognitive processing capacity at any given time. Being an asynchronous resource, it gives students the flexibility and autonomy to work through the resource independently without requiring any training from instructors. In line with Mayer’s Pre-Training principle, an ‘Orientation’ page is included at the start to provide learners simple guidance on how to navigate through the resource (Corbeil et al. 2021). This is followed by a short pre-course quiz to test learners’ existing understanding of Kawasaki Disease and to identify gaps in their knowledge. The resource takes a systematic approach by taking the learner through key aspects of the condition such as epidemiology, aetiology, diagnostic criteria and treatment.

Interactive activities are weaved into the resource to consolidate learning and to keep learners engaged; an example of this is the ‘drag and drop’ exercise as shown in Figure 1. The resource concludes with a self-assessment and an evaluation where learners are given the opportunity to provide feedback.

**Figure 1.**
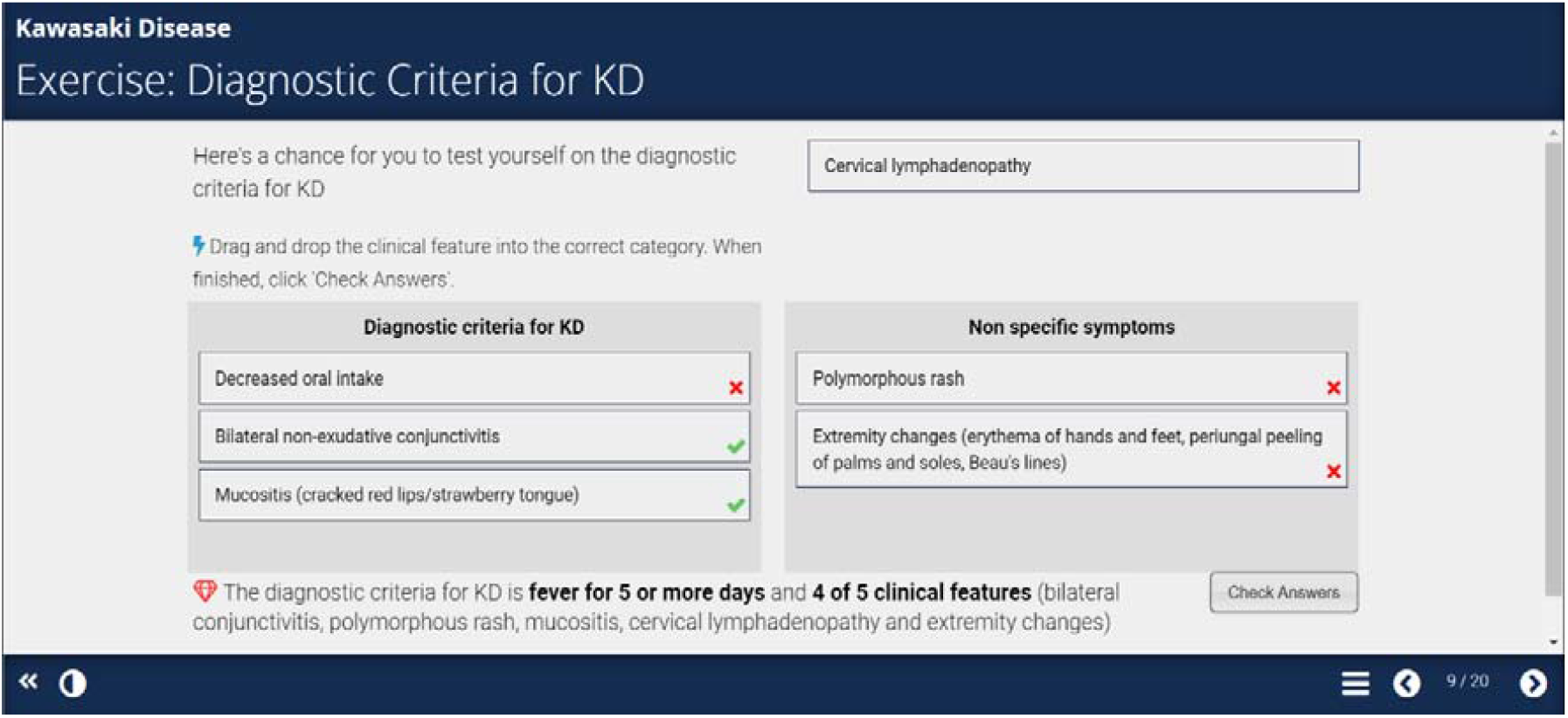
An example of an interactive activity

The Coherence Principle states that all non-essential and extraneous material should be eliminated as this can burden the cognitive load. While colourful images and sound effects can be attention-grabbing and appealing, research from controlled laboratory settings has found that excluding unnecessary graphics and text leads to greatest knowledge transfer and retention (Muller et al. 2008). The application of this principle is demonstrated in Figure 2, where only salient clinical points on ‘Incomplete Kawasaki Disease’ are included.

**Figure 2.**
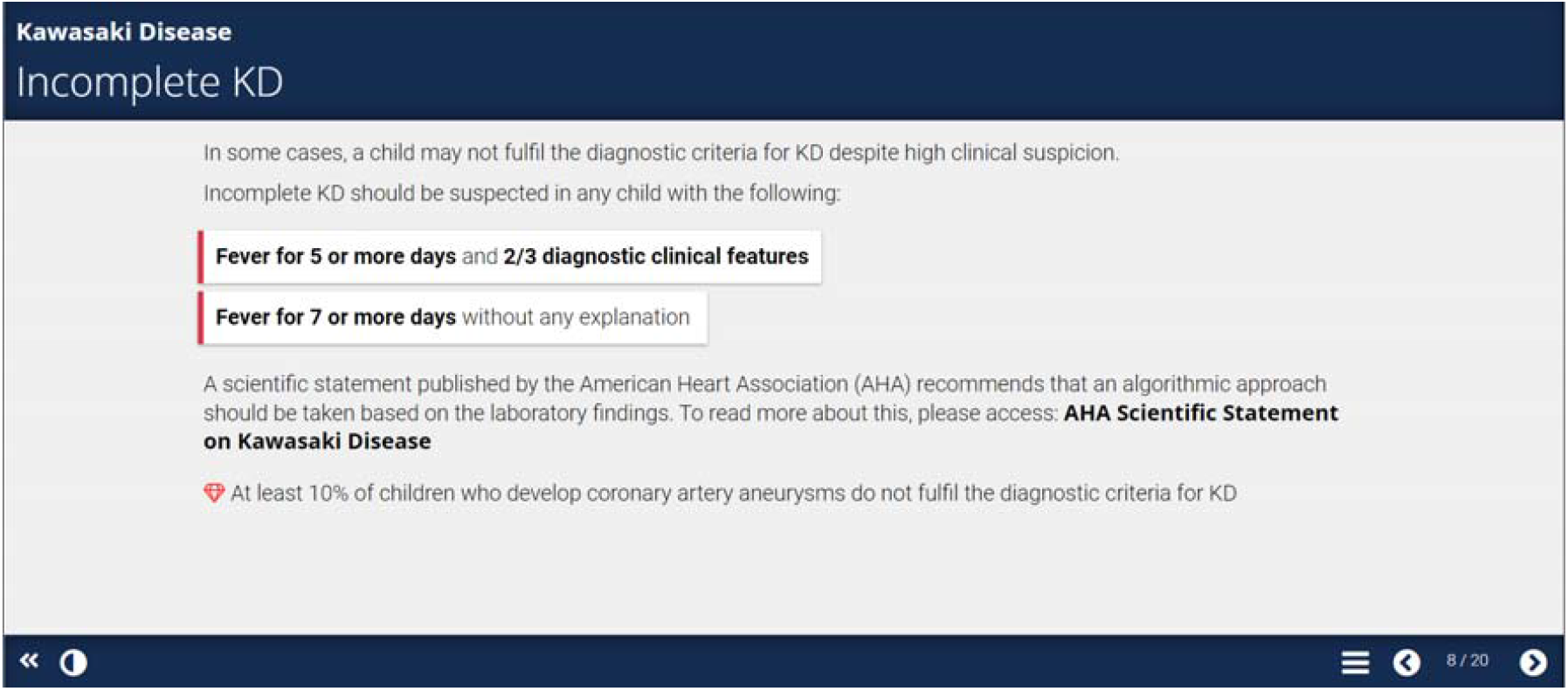
Coherence Principle applied in the resource

The use of relevant pictures with words can enhance learning and lead to greater learner recall as demonstrated by Mayer’s Multimedia Principle (Fletcher and Tobias 2005; Mayer 2010). This is demonstrated in Figure 3, where it shows a diagram of a young boy with the diagnostic clinical features of Kawasaki Disease. Learners interact with the resource by clicking the arrow beneath the diagram, where a more in-depth description of each clinical feature is displayed with the corresponding abnormality lighting up on the body of the child.

**Figure 3.**
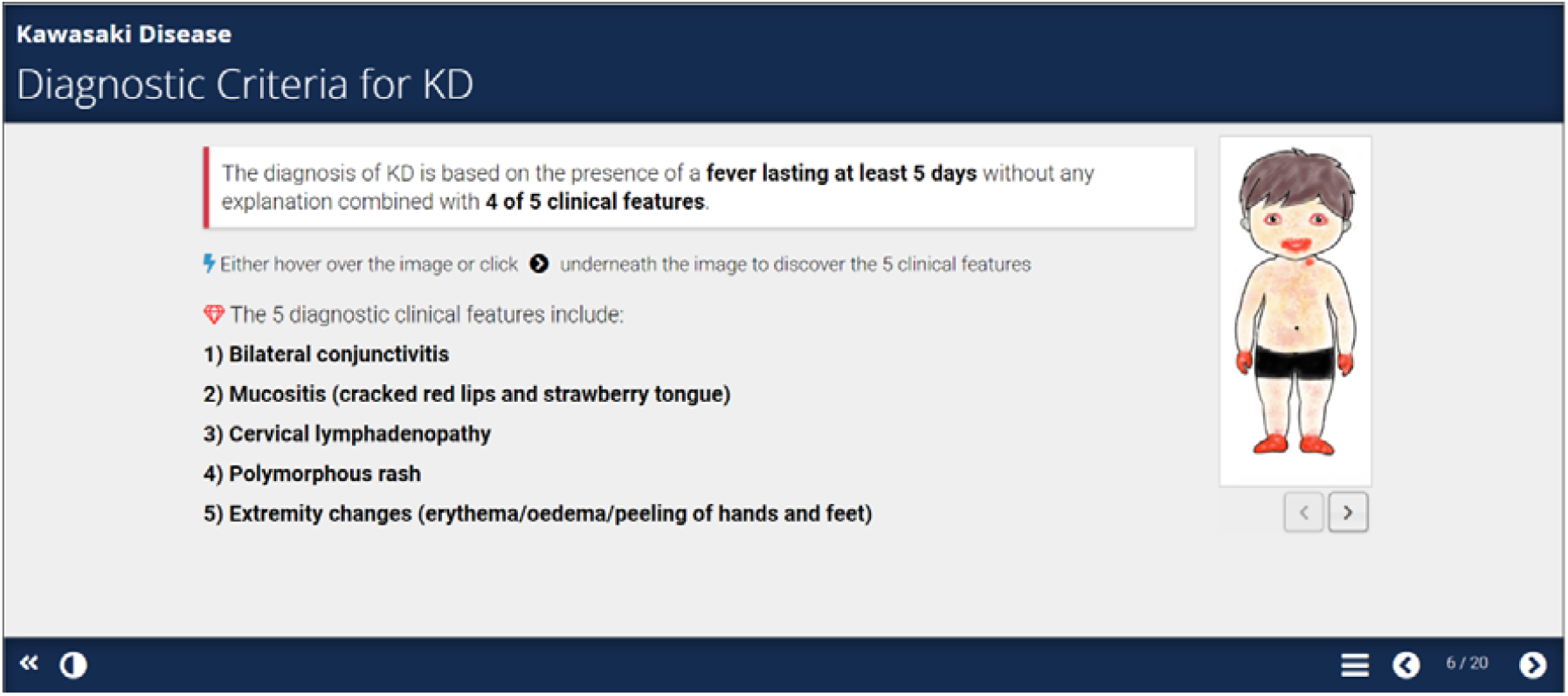
Application of Multimedia Principle

In the implementation stage, the Xerte link was sent via email to a group of medical students of all year groups who had expressed prior interest in undertaking the e-learning tutorial.

Following this, a summative evaluation was carried out on a small group of learners.

### Preliminary Evaluation

An evaluation form was included at the end of the e-learning resource for the purpose of a summative evaluation. Only five questions were included in the survey (see Box 2) in keeping with research that has found that shorter surveys yield greater response and completion rates (Kost and de Rosa 2018). The questions were based on Zaharias and Poylymenakou’s (2009) usability evaluation method, which demonstrates reliability and validity. This method goes beyond merely the cognitive and task-oriented paradigm by focusing on the holistic experience of the user by factoring in affective-emotional aspects of interaction.

Questions 1-2 are based on instructional design parameters of ‘navigation’ and ‘visual design’ while questions 3-5 fall into the category of ‘intrinsic motivation to learn’, which can be characterised as an inner drive to carry out an activity, of which the reward is enjoyment from the activity itself (Davis and Csíkszentmihályi 1977).

#### Box 2

Survey questions

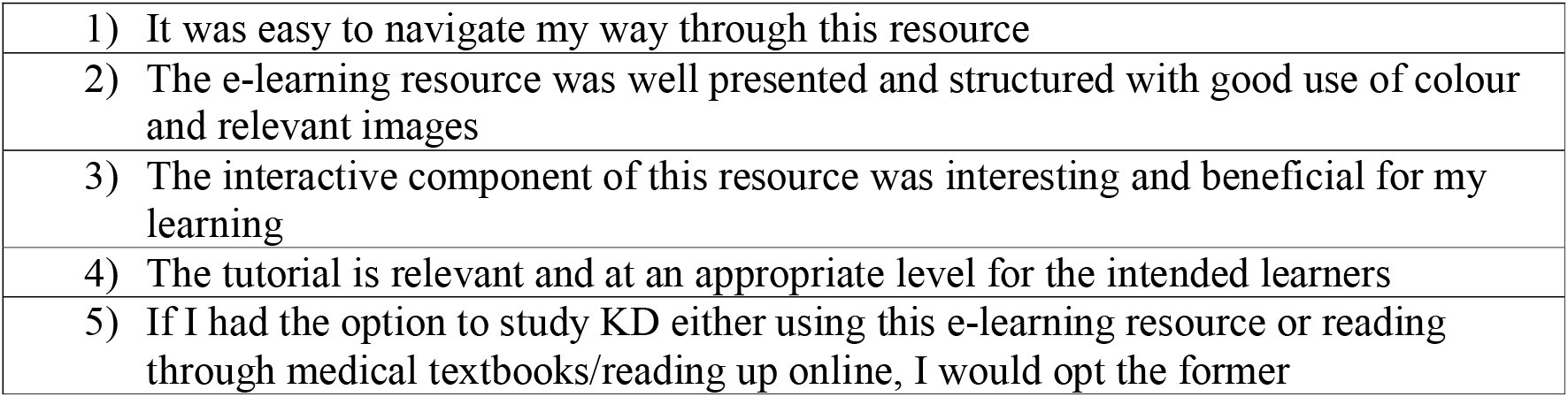

## Results

With research showing that 95% of usability problems can be identified in a sample of five to six people (Nielsen 1994), a sample size of seven is considered adequate, although a bigger sample size would be required in evaluation research. The feedback was overwhelmingly positive with all undergraduate learners agreeing or strongly agreeing with all five statements (see Figure 4). The feedback provided was anonymous, therefore the demographics of the responders and their current level of study is unknown. The comments left in the free-text mainly related to the interactivity and visual aspect of the resource such as the ‘drag and drop’ feature being unclear and suggestions for more pictures to be included in the slides.

**Figure 4.**
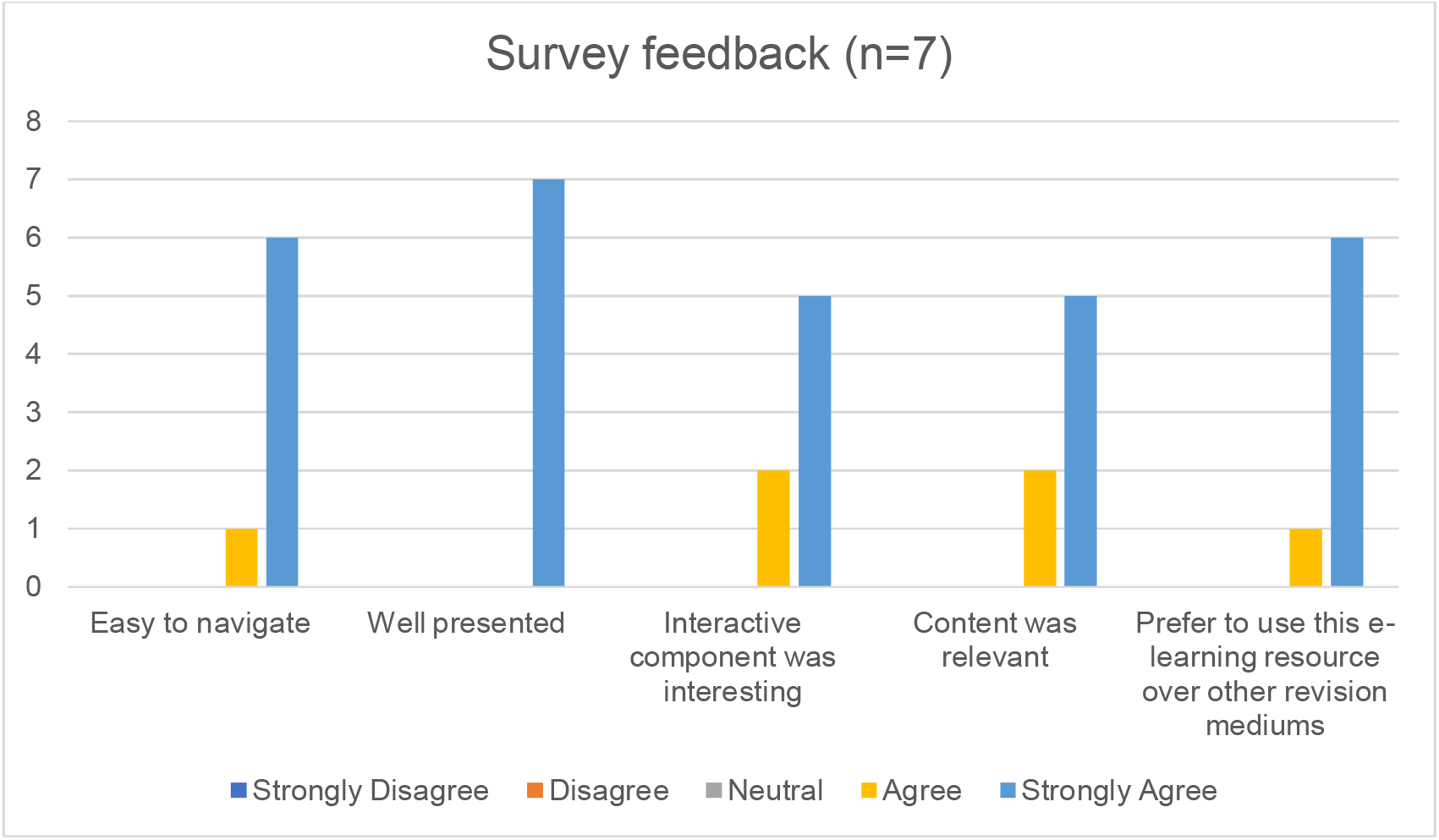
Results of student feedback

Whilst the results of this preliminary evaluation are encouraging, it is important to note that there are some limitations to this small-scale evaluation in that the questions only address a few cherry-picked surface level aspects of this e-learning resource. This raises the question of whether medical educators may consciously, or subconsciously, be selective in the areas of the e-learning resource they evaluate in order to gain favourable outcomes. Indeed, it has long since been recognised that e-learning resources suffer from poor regulation of content validity, reliability and credibility, thus this calls for a more in-depth and robust evaluation to ensure quality assurance and improvement (Leacock and Nesbit 2007).

### Evaluation protocol

At its core, evaluation refers to making a value judgment or determining the worth or merit of something (Worthen et al. 1990; Cook 2010). In the context of digital education resources, it is essential that medical educators are creating resources that are educationally beneficial and meaningful for learners (Zehry et al. 2011). Thus, further development of the digital educational tool largely depends on the effectiveness of the evaluation process.

Whilst it is recognised that effective evaluation comes with significant challenges and can be complex (Leacock and Nesbit 2007), given the nature and purpose of this resource, this should not be the case. More specifically, this is a relatively low-stake educational tool, designed to aid revision. Its aim is to supplement, not replace, existing educational activities within the curriculum (e.g., clinical placement, lectures, books). Thus, evaluation should not be over complicated or resource intensive. Essentially it is important to evaluate whether the resource is effective in meeting its intended outcomes, whether it meets learners’ needs, and to establish the suitability of the educational context and pedagogy. According to Lai and Bower (2019), evaluation of learning technologies typically focus on learning, affective elements, behaviour, technology, design, teaching/pedagogy, presence and institutional environment, and these evaluation themes will be considered in future work.

1. *Evaluating the ability of the resource to meet its intended outcomes* - Evaluation of learning will be achieved via a pre- and post-knowledge test, put together by the resource developers in consultation with subject experts. Indeed, evaluation and further development must take into account not just the learners’ opinions and perspectives, but that of other stakeholders such as medical educators, expert clinicians and technology experts (Cook 2010). Affective elements will be examined through measures of self-efficacy (Schunk (1996), confidence and intrinsic motivation (Zaharias and Poulymenakou (2009). These are well understood psychological constructs that can easily be measured through pre-existing measures, adapted for the context. For example, Keller’s (1993) Instructional Material Motivation Survey and Motivated Strategies for Learning Questionnaire (Pintrich 1991) have been used in previous studies. Behaviour will be evaluated by asking learners how and when they have used the resource through a range of open-ended questions.
2. *Evaluating the ability of the resource to meet learners’ needs* – The resource is designed with undergraduate students in mind at all levels of study. However, it is unclear, especially as the demographics of the responders who completed the preliminary evaluation is unknown, rendering possible overrepresentation of certain year groups. Further evaluation will therefore consider the views of other stakeholders regarding the suitability of learning outcomes, content, and activities and if they meet the needs of learners at all levels of study, or whether it may be more suitable for learners who have already attended a clinical placement in paediatrics. Further revisions of the resource may need to reflect the spiral nature of the medical curriculum.
3. *Evaluating the educational context* - The educational context is digital and for this reason evaluation will focus on criteria related to design, content, usability and functionality. To this end, evaluation will be more comprehensive, yet straightforward by using pre-existing models such as the Learning Object Review Instrument (LORI), which comprises of nine items: content quality, learning goal alignment, feedback and adaptation, motivation, presentation design, interaction usability, accessibility, reusability and standards compliance. It must be mentioned that the dimensions LORI is founded on can be interpreted broadly and thus allows a fine balance to be struck between depth of assessment and time; an important consideration among busy clinicians (Leacock and Nesbit 2007).While ADDIE was used in a linear fashion in the production of this resource, some favour a more iterative approach by placing the evaluation component at the centre of the model and collecting regular formative evaluation after each step in the process. An advantage of this is that it allows the designer to identify strengths and weaknesses and make amendments throughout the process, which can lead to continuous improvement of the educational tool (Allen 2006). Alternatively, the resource can be rolled out provisionally in a pilot implementation in the fourth step of the process, where formative evaluation can identify shortcomings of the resource for further analysis and revision to take place (Welty 2008). Following this would be a final implementation where a summative evaluation is undertaken, and conclusions may be drawn on various elements of the resource. It is important to note that there is no standard number of formative evaluations that should take place; rather this depends on multiple factors such as the budget and time available, as well as the circumstances surrounding the project (Welty 2008).

### Summary

In light of rising popularity in the use of digital learning resources in the teaching of undergraduate medical students, it is imperative that medical educators ensure that evaluation is robust and that it is given the same amount of thought and attention as the earlier stages of the instructional process. ADDIE is a useful framework that can be used to inform instructional design among novice content creators, and while some choose to take an A-to-E approach concluding with a summative evaluation, the benefits of taking an iterative approach with regular formative evaluation has been well documented in literature (Allen 2006). Carrying out a mixed qualitative and quantitative analysis on specific components of evaluation that are pertinent to the resource may be effective, while utilising established evaluation instruments such as LORI to evaluate the technological aspects of the resource such as content quality, presentation design and usability may ensure quality control (Leacock and Nesbit 2007).

## Data Availability

All data produced in the present work are contained in the manuscript.

